# The use of germicidal ultraviolet light, vaporised hydrogen peroxide and dry heat to decontaminate face masks and filtering respirators contaminated with a SARS-CoV-2 surrogate virus

**DOI:** 10.1101/2020.06.02.20119834

**Authors:** Louisa F. Ludwig-Begall, Constance Wielick, Lorène Dams, Hans Nauwynck, Pierre-Francois Demeuldre, Aurore Napp, Jan Laperre, Eric Haubruge, Etienne Thiry

## Abstract

**Background:** In the context of the ongoing severe acute respiratory syndrome coronavirus 2 (SARS-CoV-2) pandemic, the supply of personal protective equipment remains under severe strain. To address this issue, re-use of surgical face masks and filtering facepiece respirators has been recommended; prior decontamination is paramount to their re-use.

**Aim:** We aim to provide information on the effects of three decontamination procedures on porcine respiratory coronavirus (PRCV)-contaminated masks and respirators, presenting a stable model for infectious coronavirus decontamination of these typically single-use-only products.

**Methods:** Surgical masks and filtering facepiece respirator coupons and straps were inoculated with infectious PRCV and submitted to three decontamination treatments, UV irradiation, vaporised H_2_O_2_, and dry heat treatment. Viruses were recovered from sample materials and viral titres were measured in swine testicle cells.

**Findings:** UV irradiation, vaporised H_2_O_2_ and dry heat reduced infectious PRCV by more than three orders of magnitude on mask and respirator coupons and rendered it undetectable in all decontamination assays.

**Conclusion:** This is the first description of stable disinfection of face masks and filtering facepiece respirators contaminated with an infectious SARS-CoV-2 surrogate using UV irradiation, vaporised H_2_O_2_ and dry heat treatment. The three methods permit demonstration of a loss of infectivity by more than three orders of magnitude of an infectious coronavirus in line with the FDA policy regarding face masks and respirators. It presents advantages of uncomplicated manipulation and utilisation in a BSL2 facility, therefore being easily adaptable to other respirator and mask types.

## INTRODUCTION

In the context of the ongoing severe acute respiratory syndrome coronavirus 2 (SARS-CoV-2) pandemic, the supply of personal protective equipment (PPE) remains under severe strain and both availability and affordability of items can be subject to fluctuations and disruptions within health-, social care and other essential public facilities. Access to PPE for the health workforce in all services (public and private, community and hospital) has been identified as a key factor in strengthening the international health system response to SARS-CoV-2, the causative agent of COVID-19 [1]; use of medical/surgical face masks and filtering facepiece respirators (FFR i.e. N95 or FFP2 or FFP3 standard or equivalent with a minimum filtration efficiency of 95% for 0.3 μm (aerodynamic mass mean diameter) of sodium chloride aerosols) has been recommended in conjunction with other mitigating measures to prevent transmission of this and other respiratory pathogens [2,3].

Since the surging global demand cannot currently be met solely by the limited capacities of expanding PPE production, national and internationally-coordinated efforts are increasingly focused on re-use of various items [4,5]. While prior decontamination is paramount to re-use of surgical masks or respirators, little information exists on effective decontamination of these typically single-use-only products. Without compromising the fit and filtration integrity of the masks themselves, decontaminating procedures must guarantee the complete inactivation of contaminating pathogens; the US Food and Drug Administration Enforcement Policy for Face Masks and Respirators issued in April 2020 recommends a robust proof of infectious bioburden reduction of three orders of magnitude for viral pathogens, specifically coronaviruses, and six orders of magnitude for either mycobacteria or bacterial spores [6].

Amongst a variety of different methods under investigation, vaporisation of the oxidizing agent hydrogen peroxide (H_2_O_2_), already a standard hospital sterilisation technology due to its broad antimicrobial activity and efficacy in surface decontamination [7], has garnered attention as a cost-effective and practical option for mask decontamination [5,6,8]. Mask shape and fit of unused 3M FFP2 NR D face masks (type 8822) were shown to remain intact subsequent to two cycles of vaporised H_2_O_2_ low pressure gas sterilisation in a study by the Dutch National Institute for Public Health and the Environment [5]; further studies have since demonstrated integrity of other respirator models after numerous cycles of H_2_O_2_ treatment and have shown virucidal and bactericidal activity of the method on a number of respiratory pathogens and/or biological indicators [8,9]. To our knowledge, no study has yet reported the effect of H_2_O_2_ treatment on SARS-CoV-2, although vaporised H_2_O_2_ decontamination trials of SARS-CoV-2 inoculated masks are reportedly underway [8].

As alternatives to chemical vaporised H_2_O_2_, two physical decontamination methods, the application of dry heat and ultraviolet (UV) irradiation, show promise for decontamination of SARS-CoV-2 contaminated masks in various settings. Heat treatment acts via denaturation of protein secondary structures thereby altering conformation of viral proteins involved in attachment to and replication within host cells and has long been recognised as an efficient method of virus inactivation [10]. Temperatures of over 65°C have previously been shown to inactivate SARS-CoV in suspension [11]; more recently, dry heat treatment of 70°C was identified to inactivate SARS-CoV-2 in solution [12] and was shown to not significantly alter filtration efficiency of N95 respirators within 20 cycles of application [13]. Easily scalable, dry heat allows mass treatment of large sample sizes and thus potentially presents a fast and efficient decontamination alternative to vaporised H_2_O_2_ in decentralised hospital centres or industrial settings. Ultraviolet germicidal irradiation with a highly energetic short-wave (254 nm) acts via viral disruption of viral DNA or RNA and constitutes a physical surface treatment for contaminated masks or FFRs. A useful sterilisation technique in a variety of applications, UV irradiation has been implemented to effectively decontaminate influenzavirus (H5N1) from two different models of FFRs (3M models 1860s and 1870) [14] and does not degrade respirator performance even after multiple applications [13,15]. It is a promising option for rapid decontamination of smaller, individual sample contingents and is as such easily adaptable to point-of-care applications. A possible concern relates to UV penetration depth, necessitating studies that not only investigate viral inactivation of decontaminated surfaces but that address a potential worst-case scenario in which viruses penetrate deeper layers of contaminated face masks and FFRs [13].

Since the utilisation, concentration and cultivation of infectious SARS-CoV-2 necessary for analyses investigating its inactivation, pose obvious problems in terms of the availability and equipping of BSL3 facilities, the use of conservative surrogates to test decontamination efficacy of various methods is justified and crucial to gain an as accurate as possible insight into SARS-CoV-2 decontamination. A recent (non-peer reviewed) publication describes inactivation of murine hepatitis virus (MHV), a SARS-CoV-2 surrogate of the same *Betacoronavirus* genus, via vaporised H_2_O_2_, heat treatment, UV exposure and other decontamination methods on FFRs; regrettably, the limited dynamic range of the MHV model (at most 10^1^-10^2^ inactivation) fell short of demonstrating a 10^3^ reduction [16].

In the present investigation into filtering facepiece respirator and surgical mask decontamination via UV irradiation, H_2_O_2_ and application of dry heat, we implemented porcine respiratory coronavirus (PRCV), a spike gene deletion mutant of transmissible gastroenteritis virus (TGEV) and a member of the *Alphacoronavirus 1* species [17,18], as SARS-CoV-2 surrogate. While PRCV, which infects the respiratory tract of swine [19], is not in the same genus as SARS-CoV-2, the two members of the subfamily *Coronavirinae* in the family *Coronaviridae* show sufficient similarities as to genome length and virion structure (notwithstanding differences in envelope glycoproteins), for them to be expected to behave similarly outside their hosts. Indeed, TGEV has previously been utilised as a surrogate for another *Betacoronavirus*, SARS-CoV, in studies investigating the persistence of coronaviruses on inanimate surfaces and/or their inactivation with biocidal agents [20,21]. The PRCV model thus combines the advantages of sufficient genetic and structural relatedness to SARS-CoV-2, the fact that PRCV can be readily propagated and assayed *in vitro* to high titres in the swine testicle (ST) cell line, and the absence of human infection risk.

This is the first description of stable disinfection of FFRs and surgical masks contaminated with an infectious SARS-CoV-2 surrogate using UV irradiation, vaporised H_2_O_2_ and dry heat treatment. The three methods permit demonstration of a loss of infectivity by more than three orders of magnitude of an infectious coronavirus in line with the FDA policy regarding face masks and respirators. It presents advantages of uncomplicated manipulation and utilisation in a BSL2 facility, therefore being easily adaptable to other respirator and mask types to which any of the three or other decontamination methods may be applied.

## METHODS

### Virus and cells

PRCV strain 91V44 [22] was passaged three times on confluent cell monolayers of the ST continuous cell line. A virus stock with a titre of 10^7.8^ TCID_50_/ml was used.

### Surgical masks and filtering facepiece respirators

All FFRs and surgical masks, commonly used by the health care community in Belgium at the time of the study, were supplied by the Department of the Hospital Pharmacy, University Hospital Centre of Liege (Sart-Tilman). Manufacturers (and models): KN95 FFR - Guangzhou Sunjoy Auto Supplies CO. LTD, Guangdong, China (2020 N°26202002240270); surgical mask (Type II) - Hangzhou Sunten Textile Co., LTD, Hangzhou, China (SuninCare™, Protect Plus). Surgical masks and FFRs were verified to be from the same respective manufacturing lot to minimise any lot-to-lot variation and to ensure consistency during future respirability and filtration performance testing. FFR and mask materials were verified via scanning electron microscopy and Fourier-transform infrared spectroscopy (spectral data obtained by direct μ-ATR-FTIR analysis (Nicolet 6700) and compared for best fit against a Hummel library of know infrared spectra of polymers as well as a Centexbel-curated spectral library). FFRs consisted of four layers of polypropylene, specifically two outer spunbound structures, an intermediate meltblown layer, and an inner spunbound layer (Supplementary Figure 3). Surgical masks were composed of three layers of polypropylene, with an outer and inner layer of spunbound polypropylene encasing a meltblown polypropylene barrier (Supplementary Figure 4).

### Inoculation of surgical masks and filtering facepiece respirators with porcine respiratory coronavirus (PRCV)

Efficacy of three different decontamination methods in inactivating an infectious coronavirus was assessed using surgical masks and FFRs experimentally inoculated with PRCV. Per decontamination method and mask type, one negative control mask or respirator (not contaminated but treated), three treated masks or respirators (PRCV-contaminated and treated), and three positive controls (PRCV-contaminated but untreated), i.e. seven masks in total, were utilised.

Prior to inoculation, the masks and FFRs were first marked with a graphite pencil to enable sample identification and to outline three square areas (34 mm x 34 mm) to the left, right and centre of the masks and FFRs, corresponding to areas to be cut out post-inoculation (coupons) with a central site of inoculation (the demarcation of the coupons and inoculation sites are shown in Supplementary Figure 1). Utilising an ultra-fine insulin syringe and needle (BD Medical), 100 μl of undiluted viral suspension in Minimum Essential Medium (MEM) were injected at the centre of each of the three square coupons under the first outer layer of mask or FFR (this to simulate a “worst case scenario” for viral inactivation in which a given decontamination method must reach a virus that has penetrated beyond the surface layer of a mask owing to respiration pressure gradients) for treated and positive control masks. In addition to inoculation of the *de facto* masks or respirators themselves, 100 μl of viral suspension was pipetted onto one elastic strap per contaminated surgical mask or FFR. The masks were allowed to dry for 20 minutes and were then individually packaged in appropriate containers (autoclaved empty tip boxes for respirators; sealable plastic bags for masks) ensuring a cold chain and minimal contamination before decontamination. Throughout, masks were handled only with sterile tweezers and gloves to limit bacterial or fungal contamination. Gloves were changed between handling of individual masks to avoid potential carry-over of inoculum.

### Decontamination of PRCV-inoculated surgical masks and filtering facepiece respirators

PRCV-inoculated surgical masks and FFRs were decontaminated utilising three different decontamination methods, UV irradiation, vaporised H_2_O_2_, and dry heat. Three FFRs and three surgical masks were subjected to each of the three decontamination methods. Three additional inoculated masks or respirators (positive controls) and the negative control mask or respirator remained individually packaged and cooled before and after each decontaminating treatment to account for the effect of time-dependent surface absorption and/or interaction on virus recovery.

### UV germicidal irradiation

Surgical masks and FFRs were individually irradiated using a LS-AT-M1 (LASEA Company, Sart Tilman, Belgium) equipped with 4 UV-C lamps of 5.5W (@UV-C). Hung vertically on a metal frame, masks and FFRs were inserted into a safety enclosure. A 2 min UV-C treatment (surgical masks) led to a fluence of 2.6J/cm^2^ per mask (1.3J/cm^2^ per side), 4 min UV-C irradiation (FFRs) led to a fluence of 5.2J/cm2 per mask. Power and irradiation time (120 s) were monitored and recorded throughout. Following irradiation, surgical masks and FFRs were unloaded and placed in individual bags.

### Vaporised H_2_0_2_

Surgical masks, FFRs and a chemical indicator were placed in individual Mylar/Tyvek pouches. Vaporous hydrogen peroxide (VHP) treatment was performed with the V-PRO Max Sterilizer (Steris, Mentor, OH) which uses 59% liquid H_2_O_2_ to generate hydrogen peroxide vapor. A 28-minute non lumen cycle was used, consisting of 2 min 40 sec conditioning (5 g/min), 19 min 47 sec decontamination (2.2 g/min) and 7 min 46 sec aeration. Peak VHP concentration was 750 ppm.

### Dry heat

Surgical masks and FFRs, hung horizontally on a metal frame, were inserted into an electrically heated vessel (M-Steryl, AMB Ecosteryl Company, Mons, Belgium) for treatment with temperatures of 102°C (± 4°C) for 60 min (± 15 min). Temperatures inside the heated vessel were recorded throughout to ensure correct exposure conditions. After termination of the treatment cycle, masks and FFRs were allowed to cool and then bagged individually.

### Elution of PRCV from decontaminated and untreated surgical masks and filtering facepiece respirators

Upon completed decontamination, the three previously determined square coupons with the focal point of PRCV inoculation at their centres were cut from the masks; the inoculated elastic strap was severed from the masks in its entirety (Supplementary Figure 2). Thus, per mask, three coupons and one elastic strap were sampled. The different layers of the coupons were separated to facilitate viral recovery and the separated layers of each individual coupon were placed together in a 15 mL Falcon tube containing 4 mL elution medium (Eagle’s MEM (Sigma)) supplemented with 2 % of an association of penicillin (5000 SI units/mL) and streptomycin (5 mg/mL) (PS, Sigma)). Pilot experiments (results not reported) indicated a cytotoxicity of eluate from H_2_O_2_-treated masks. To combat this effect, presumably due to residual H_2_O_2_ in the suspension, the elution medium was supplemented with 20% FCS and 0.1% β-mercaptoethanol in a total volume of 4 mL for virus elution subsequent to vaporised H_2_O_2_ treatment. Sterile sets of scissors and tweezers were used for each coupon, thus avoiding cross-contamination between inoculation points. The mask and respirator coupon layers and elution medium were mixed for 20 minutes at maximum speed (2500 rounds per minute (rpm)) using a multitube vortex mixer (VWR VX-2500 Multi-Tube Vortexer). Supernatant eluates were recovered via pipette and either directly utilised in downstream applications or stored at - 80°C until further analysis.

### Quantification of infectious PRCV eluted from decontaminated and untreated surgical masks and filtering facepiece respirators

Titres of infectious PRCV recovered from individual coupons and straps of decontaminated surgical masks and respirators were determined separately using a TCID_50_ assay in ST cells. Briefly, tenfold dilutions were made from each sample and 50μl of each dilution were inoculated in each of four wells of a 96-well plate. After one hour of incubation, 100μl of medium (MEM supplemented with 10% FCS and antibiotics) were added to each well. Due to toxicity in the undiluted inoculum, the inoculum was completely removed and 150μl of medium were added. Four days after inoculation, monolayers were analysed for the presence of cytopathic effect by light microscopy. Virus titres were calculated using the Reed and Muench method [23]. Back titrations of virus inoculum stocks were performed in parallel to each series of decontamination experiments.

### Data analysis and statistics

Statistical analyses of differences in infectious viral titres were performed using GraphPad Prism 7 (Graph-Pad Software) and P-values were computed by using a two-sided independent sample t-test, where ****P<0.0001, ***P<0.001, **P<0.01, *P<0.05, and ns is P≥0.05.

## RESULTS

Back titrations of virus inoculums performed in parallel to each series of experiments confirmed PRCV inoculum titres to be within a range of 6.31×10^6^ to 2×10^7^ TCID_50_/mL for all experiments.

The cell culture limit of detection (LOD) was 6.31×10^0^ TCID_50_/mL for all assays. An initially observed H_2_O_2_ cytotoxicity and correspondingly elevated LOD of 6.31×10^1^ TCID_50_/mL of H_2_O_2_-treated coupon eluates was corrected via β-mercaptoethanol and FCS supplementation of elution medium; elevated cytotoxicity of H_2_O_2_-treated strap eluates (SM and FFR) could not be neutralised and remained at 6.31×10^1^ TCID_50_/mL. Values below the LOD were considered as ≤6.31×10^0^ TCID_50_/mL, with the exception of measurements concerning H_2_O_2_-treated straps.

### Infectious PRCV is recovered at high titres from untreated surgical mask- and filtering facepiece respirator coupons, at lower titres from surgical mask straps, and remains under the limit of detection following recovery from filtering facepiece respirator straps

Recovery of infectious PRCV from inoculated untreated surgical masks and FFRs was analysed in ST cells. Comparable high levels of infectious virus were recovered from PRCV-inoculated, untreated left, right and middle coupons of all surgical masks. Mean values were 2.83×10^5^ (±2.0) TCID_50_/mL, 1.69×10^5^ (±1.81) TCID_50_/mL and 4.25×10^5^ (±2.88) TCID_50_/mL for recovery from positive control coupons of the UV, H_2_O_2_ and dry heat assays, respectively. Mean strap recovery values were also similar between experiments, however they were lower by two to three orders of magnitude than surgical mask coupon recovery values, with mean values of 5.88×10^3^ (±2.80) TCID_50_/mL, 1.21×10^2^ (±0.71) TCID_50_/mL, and 3.92×10^2^ (±4.56) TCID_50_/mL for straps utilised as positive controls in the UV, H_2_O_2_ and dry heat assays, respectively (Figure 1). Recovery from FFR coupons yielded mean infectious virus at 6.96×10^4^ (±13.27) TCID_50_/mL, 1.80×10^5^ (±1.58) TCID_50_/mL and 2.16×10^4^ (±3.28) TCID_50_/mL for positive controls of the UV, H_2_O_2_ and dry heat assays, respectively. Recovery values for infectious virus from FFR straps remained below the LOD for UV and dry heat positive controls; recovery from straps used as positive control in the H_2_O_2_ experiments was 1.75 x10^3^ (±0.43) TCID_50_/mL (Figure 1).

**Figure 1.**
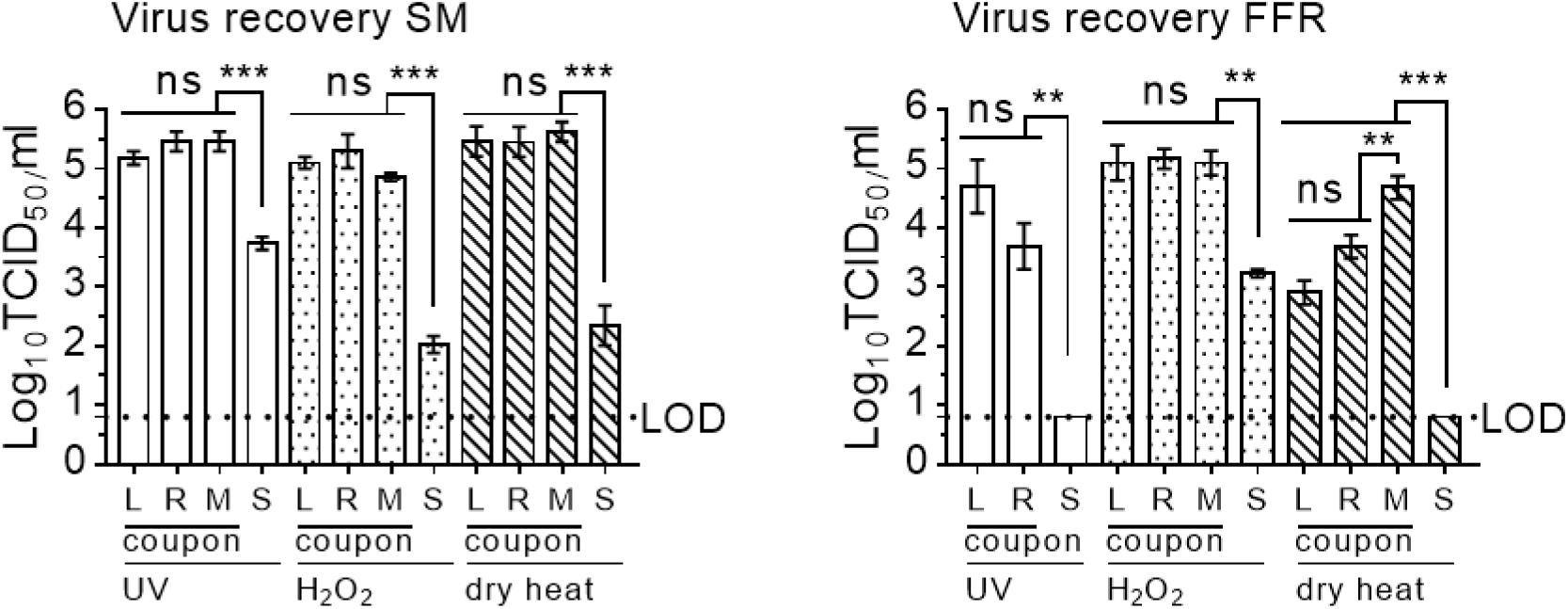
Recovery of virus after elution from inoculated, untreated surgical masks and filtering facepiece respirators. Recovery of infectious porcine respiratory coronavirus from inoculated untreated surgical masks (SM) and filtering facepiece respirators (FFR) was analysed in swine testicular cells. The cell culture limit of detection (LOD) was 0.8 log_10_ TCID_50_/mL (6.31×10^0^). Similar levels of virus recovery were detected for left, right and middle (L, R, M) coupons of masks and respirators; recovery efficacy of infectious virus from straps (S) deviated significantly in all analyses from the mean of all coupons and remained below the LOD for all assays performed on FFR straps. P-values were computed by using a two-sided independent sample t-test to calculate differences between individual coupon values and differences between mean values of all coupons and straps, where ****P<0.0001, ***P<0.001, **P<0.01, *P<0.05, and ns is P≥0.05.

### UV irradiation, vaporised H_2_O_2_ and dry heat treatment reduce infectious PRCV by more than three orders of magnitude on surgical mask and filtering facepiece respirator coupons and render it undetectable in all decontamination assays

Following UV irradiation (2 minutes exposure time), exposure to vaporised H_2_O_2_, and dry heat treatment of surgical masks, all titres for virus recovered from coupons and straps remained below the respective LOD of the assay, showing a total loss of infectivity of more than five orders of magnitude for all three treatments on coupons (2.83×10^5^ (±2.0) TCID_50_/mL, 1.69×10^5^ (±1.81) TCID_50_/mL and 4.25×10^5^ (±2.88) TCID_50_/mL, respectively); titres of virus recovered from treated surgical mask straps were reduced by three orders of magnitude post UV irradiation (5.88×10^3^ (±2.80) TCID_50_/mL), by two orders of magnitude for heat-treated straps (3.85×10^2^ (±4.56) TCID_50_/mL), and by one order of magnitude for H_2_O_2_-treated straps (5.78×10* (±7.10) TCID_50_/mL).

Decontamination treatment effects followed a similar pattern of viral inactivation for FFR coupons decontaminated via H_2_O_2_ and dry heat, reducing viral titres by over five orders of magnitude (1.80×10^5^ (±1.58) TCID_50_/mL) and four orders of magnitude (2.16×10^4^ (±3.28) TCID_50_/mL, respectively. While UV irradiation was sufficient to deactivate PRCV on surgical masks with an exposure time of 2 minutes, it was shown to be insufficient to achieve viral inactivation by more than three orders of magnitude on a different lot of FFRs in a trial run (results not shown); a 4 minute exposure time was thus tested for FFRs, reducing viral titres by over four orders of magnitude, from 6.96×10^4^ (±13.27) TCID_50_/mL to below the LOD (Figure 2). The impact of decontamination could not be measured for UV-or dry heat-treated FFR straps due to insufficient recovery of infectious virus in positive FFR strap controls. Hydrogen peroxide treatment of FFR straps resulted in a reduction of infectious virus load by 1.69×10^3^ (±0.43) TCID_50_/mL.

**Figure 2.**
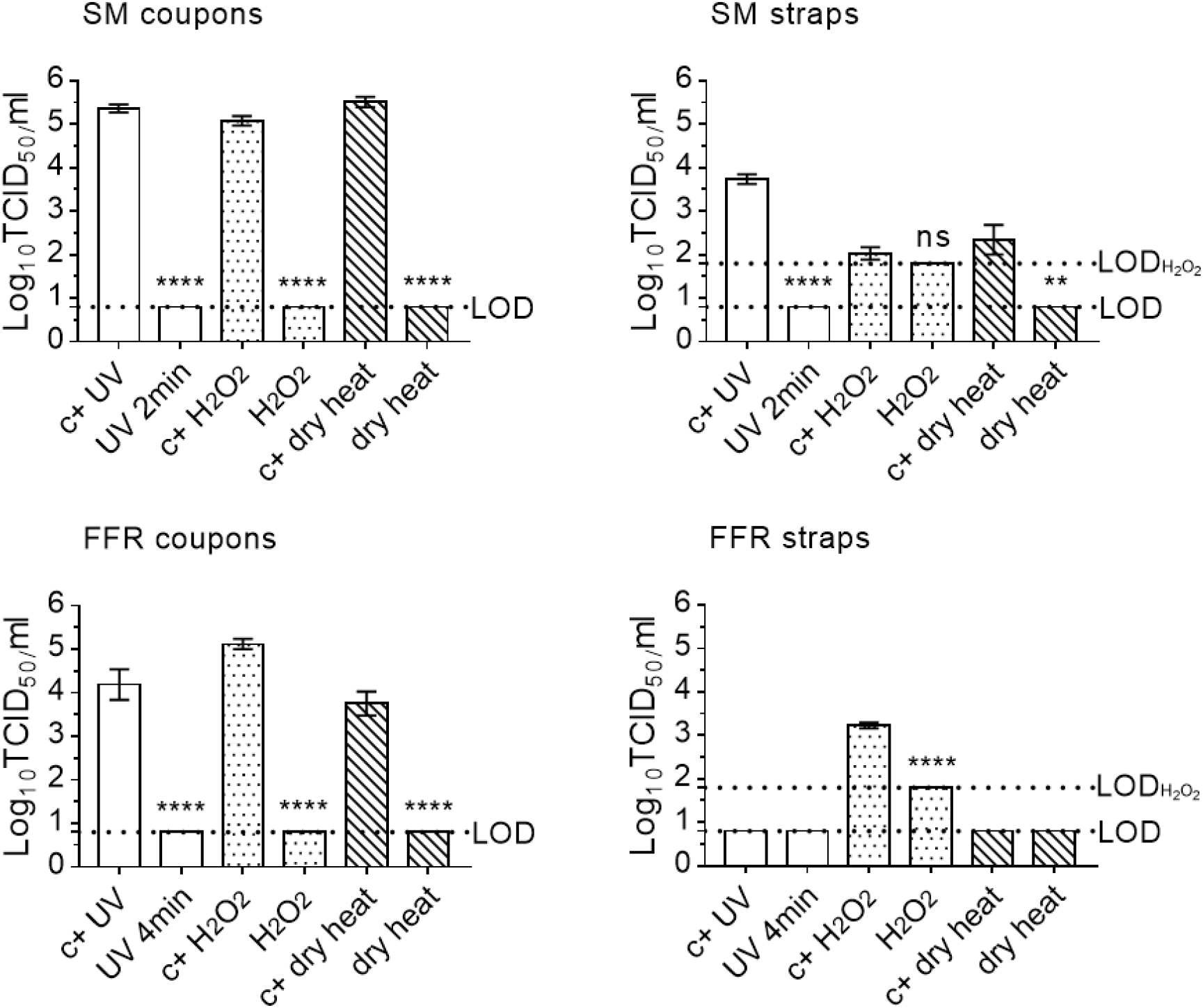
Effect of three decontaminating treatments on PRCV-inoculated surgical mask- and filtering facepiece respirator coupons and straps. The infectivity of porcine respiratory coronavirus (PRCV) recovered from surgical masks (SM) and filtering facepiece respirators (FFR) decontaminated via exposure to ultraviolet light (UV), vaporised hydrogen peroxide (H_2_O_2_), or dry heat treatment was analysed in swine testicular cells. The cell culture limit of detection (LOD) was 0.8 log_10_ TCID_50_/ml for all analyses except those concerning H_2_O_2_-treated SM or FFR straps (1.8 log_10_ TCID_50_/ml). Per decontamination method, nine PRCV-inoculated, decontaminated coupons (n=9) and three inoculated, decontaminated straps (n=3) were analysed in parallel to inoculated, untreated, positive control coupons (n=9) and straps (n=3). Sample size deviated for UV-decontaminated FFR respirators, where n=6 coupons (left and right) were analysed. P-values were computed by using a two-sided independent sample t-test, where ****P<0.0001, ***P<0.001, **P<0.01, *P<0.05, and ns is P≥0.05.

## DISCUSSION

This is, to our knowledge, the first description of stable disinfection of filtering facepiece respirators and surgical masks contaminated with an infectious SARS-CoV-2 surrogate using UV irradiation, vaporised H_2_O_2_, and dry heat treatment. While other reports have described efficacy of various decontamination methods on a range of biological indicators, few studies report on the validated decontamination of masks or respirators inoculated with SARS-CoV-2 or a conservative surrogate virus, the former being limited by the availability of BSL3 facilities, the latter by the lack of a stable high-titre model virus with an adequate dynamic range to fulfil FDA policy requirements of demonstrating a loss of infectivity by more than three orders of magnitude [8,16].

PRCV, a SARS-CoV-2 surrogate and fellow member of the *Coronavirinae* subfamily, is classed as a BSL2 pathogen. It can be cultured to high viral titres in permissive ST cells, thus possessing advantages of uncomplicated manipulation and utilisation in a BSL2 facility and a wide dynamic range. Here we demonstrate successful recovery of high quantities of infectious PRCV from inoculated, otherwise untreated surgical masks and FFR coupons, with recovery titres stably averaging over 10^5^ TCID_50_/mL for elution from mask coupons and ranging between values of over 10^3^ to 10^5^ TCID_50_/mL for elution from FFR coupons. Slightly lower recovery values of the dry heat assay are probably attributable to longer delays between inoculation and elution of infectious virus. Three decontamination methods, chemical vaporised H_2_O_2_, physical inactivation via UV irradiation and dry heat treatment, were tested for their ability to inactivate infectious PRCV on inoculated surgical masks or FFRs. All three methods rendered PRCV inoculated under the outer surface layer of mask and respirator coupons undetectable, successfully reducing the infectious load by more than three orders of magnitude.

Since carrier surfaces likely influence decontamination efficacy, we aimed to examine viral inactivation not only on the *de facto* respirators or masks themselves but on their elastic straps that may become equally contaminated. We compared titres of infectious virus recovered from inoculated, untreated straps and those inoculated and subsequently decontaminated via either UV irradiation, vaporised H_2_O_2_, or dry heat treatment. All decontamination methods rendered PRCV undetectable following recovery from straps; however, owing to insufficient virus recovery from untreated mask straps, only UV decontamination of surgical mask straps could be successfully validated as reducing viral loads by more than three orders of magnitude. Recovery of infectious virus applied to FFR straps proved impossible with a simple elution medium; however, when 20% FCS and 0.1% β-mercaptoethanol were added to the elution medium (intended to combat H_2_O_2_ cytotoxicity), infectious virus was recoverable from FFR straps. Further studies are planned to elucidate these effects, which may potentially be associated either to inherent virucidal properties of the elastic materials or attributable to poor elution from the straps. The fact that supplemented medium enabled recovery of infectious virus from FFR straps suggests that either of the supplemented constituents could have had a protective effect shielding infectious virus from potential virucidal impacts of the straps. It is worth noting that, although the simple MEM matrix used to inoculate and elute virus loads has similarities to the natural biocontamination arising from use (i.e. build-up of inorganic salts), a complex biocontamination (e.g. polymer chains in sputum) may not be replicated.

Decontaminating treatments, by their very nature, are known to have inherently detrimental side effects; particularly after multiple cycles, the integrity of decontaminated objects may be compromised. UV irradiation, vaporised H_2_O_2_, or dry heat treatment, have previously been shown to not significantly impact performance of polypropylene-based FFRs and/or masks in a number of studies [8,9,13,15]; however, others have shown that the maximum number of decontamination cycles may be limited by the respirator model and treatment conditions required for inactivation [24]. In the present investigation, masks and FFRs were effectively destroyed at the end of each cycle. Since, the safe reuse of masks and FFRs is important both in the context of the current Covid-19 pandemic and beyond, when cost-effectiveness, environmental benefits and logistic considerations will advocate a continued decontamination of these previously single-use items, further work is planned to investigate how many decontamination cycles may be safely applied to these previously single use items.

## CONCLUSIONS

In conclusion, we describe successful validation of three decontamination methods, UV irradiation, vaporised H_2_O_2_ and dry heat treatment, in inactivating an infectious coronavirus in line with the FDA policy regarding face masks and respirators. Without enough proof of inactivation, we cannot recommend safe decontamination of respirator straps and suggest additionally treating straps separately by exposure to 70% ethanol [20] until further results become available. Since H_2_O_2_ breaks down to water and oxygen, concerns for toxicity of vaporised H_2_O_2_ treatment are generally held to be very low risk [25]; however, to eliminate potential exposure of users, we recommend that a short aeration time should be respected. The PRCV surrogate supplements existing data regarding decontamination of surgical masks and FFRs, and both it and the different decontamination methods tested, are easily adaptable to other respirator and mask types, presenting a useful conservative model for stable validation of coronavirus decontamination.

## Data Availability

All data generated or analysed during this study are included in this published article.

## ACKNOWLEDGEMENTS

The authors express their sincere gratitude to Amélie Matton and Frédéric de Meulemeester (AMB Ecosteryl, Mons, Belgium), Axel Kupisiewicz (LASEA, Sart-Tilman, Belgium), Pierre Leonard (Solwalfin, Belgium) for suggestions and technical and administrative support and thank Chantal Vanmaercke and Carine Boone for their excellent technical support.

## FUNDING SOURCE

This work was supported by a grant from the Walloon Region, Belgium (Project 2010053 -2020-390 “MASK - Decontamination and reuse of surgical masks and filtering facepiece respirators”).

**Non-standard abbreviations**
FFR: filtering facepiece respirator
SM: surgical mask
SARS-CoV-2: severe acute respiratory syndrome coronavirus 2
PRCV: porcine respiratory coronavirus

## SUPPLEMENTARY FIGURES

**Supplementary Figure 1.**
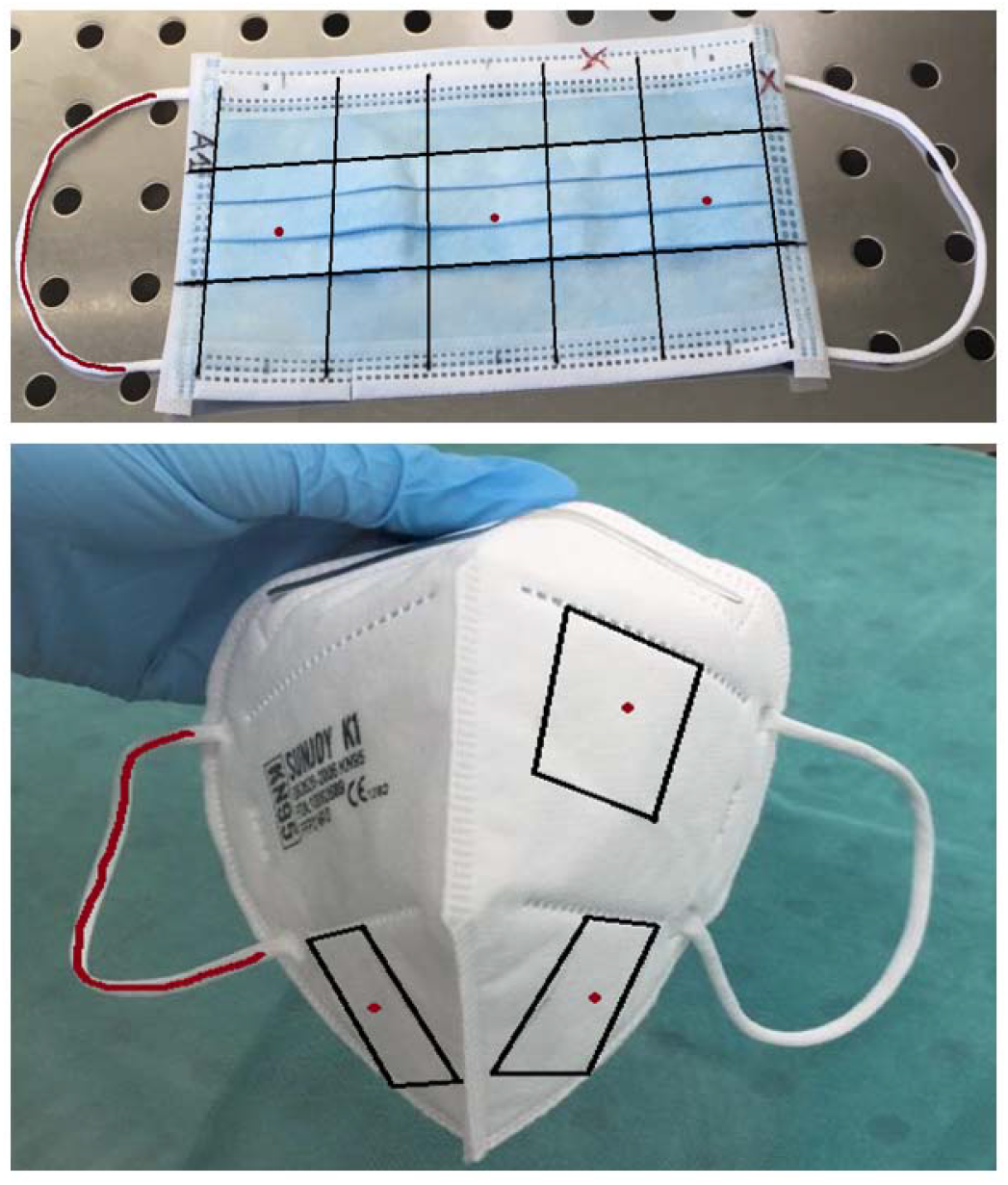
Demarcation of the three coupons (black squares), future PRCV inoculation sites (red dots), and marked elastic strap are shown on a representative surgical mask (top) and filtering facepiece respirator (bottom).

**Supplementary Figure 2.**
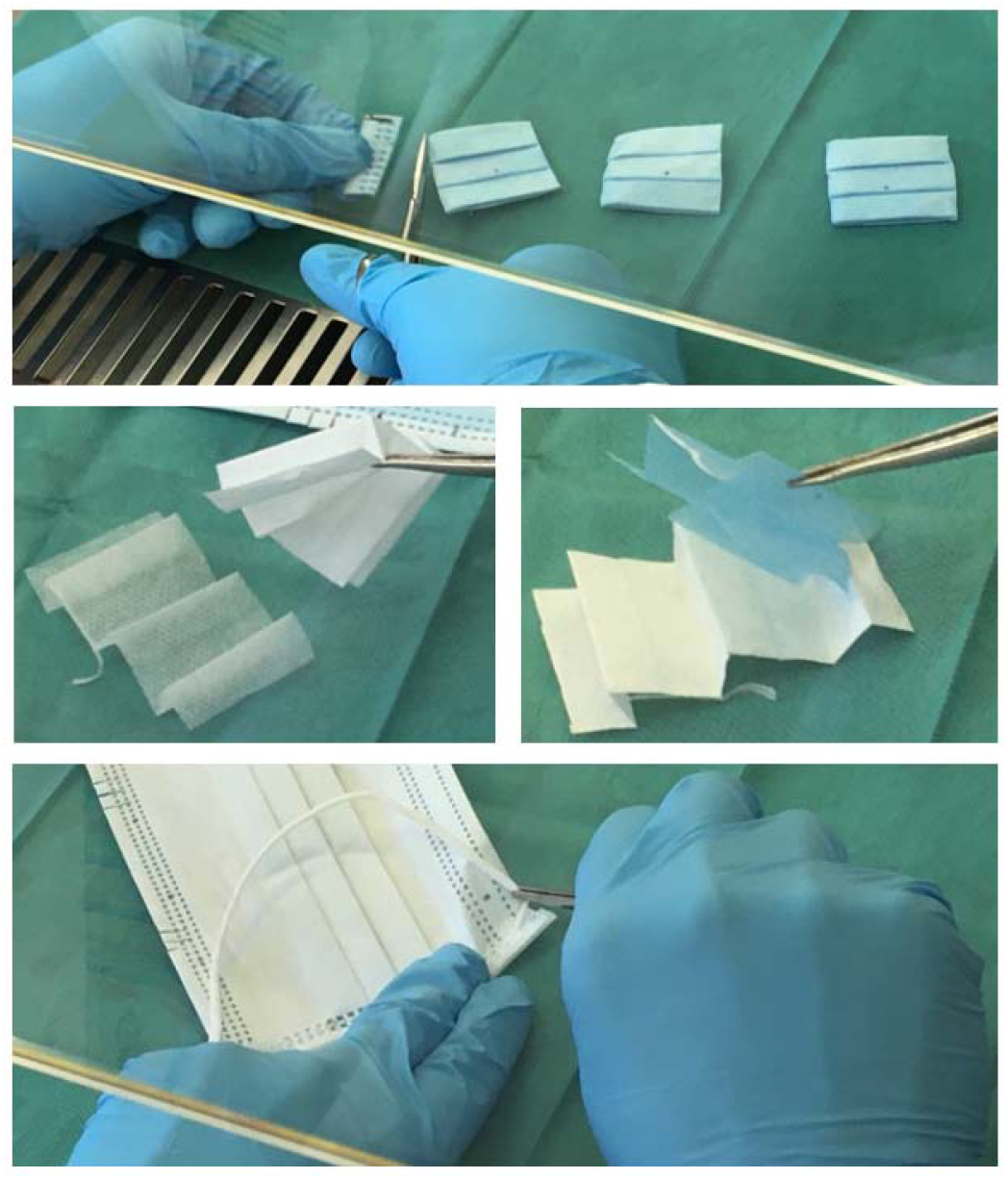
Three square coupons with a focal point of PRCV inoculation at their centres are cut from a surgical mask (top); to facilitate viral elution, different layers of the coupons are separated (middle); an inoculated elastic strap is severed from a surgical mask (bottom).

**Supplementary Figure 3.**
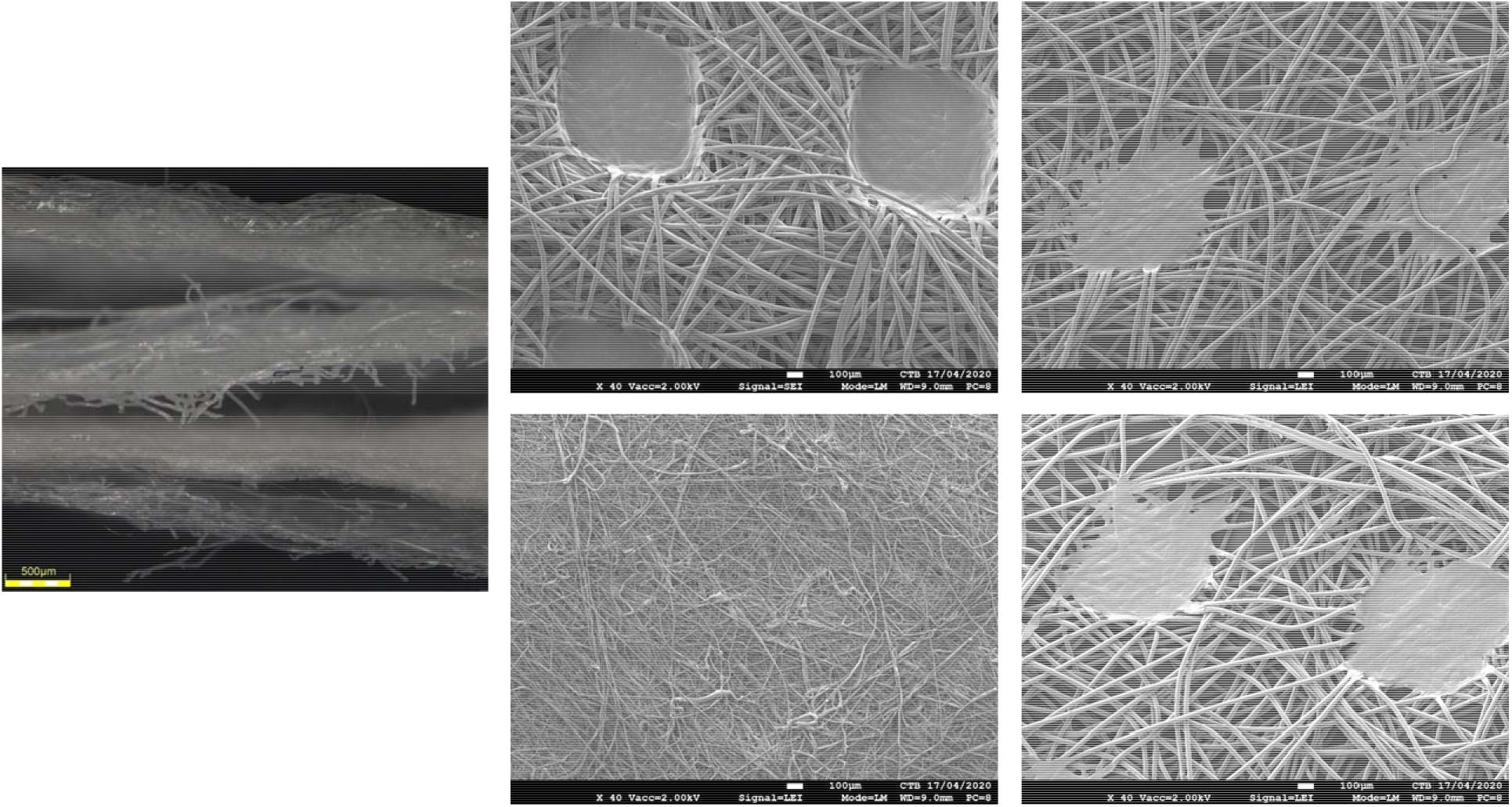
Scanning electron microscopy of a KN95 filtering facepiece respirator (FFR). Far left: Cross section view of the FFR. The two top layers are polypropylene spunbound structures, the third layer from the top is a polypropylene meltblown layer, the fourth layer is a polypropylene spunbound layer facing the wearer (the inner layer). Group of four: Details of all four polypropylene layers of the FFR. The outermost spunbound layer, the second spunbound layer, the intermediate meltblown layer, and the inner spunbound layer (61.4, 27.5, 24.4, and 28. 8 grams/m^2^ from outer to inner layer) are shown in clockwise direction. The dots in the spunbound layers show points where the fibres are melted together to ensure layer integrity. Pictures were taken on a JEOL JSM-7600F Analytical Ultrahigh resolution thermally assisted field-emission gun source scanning electron microscope at 40 times magnification. Prior to taking the pictures, samples were coated with a platinum/palladium layer (suited to non-conductive samples) using a JEOL JFC-2300 High Resolution sputter coater.

**Supplementary Figure 4.**
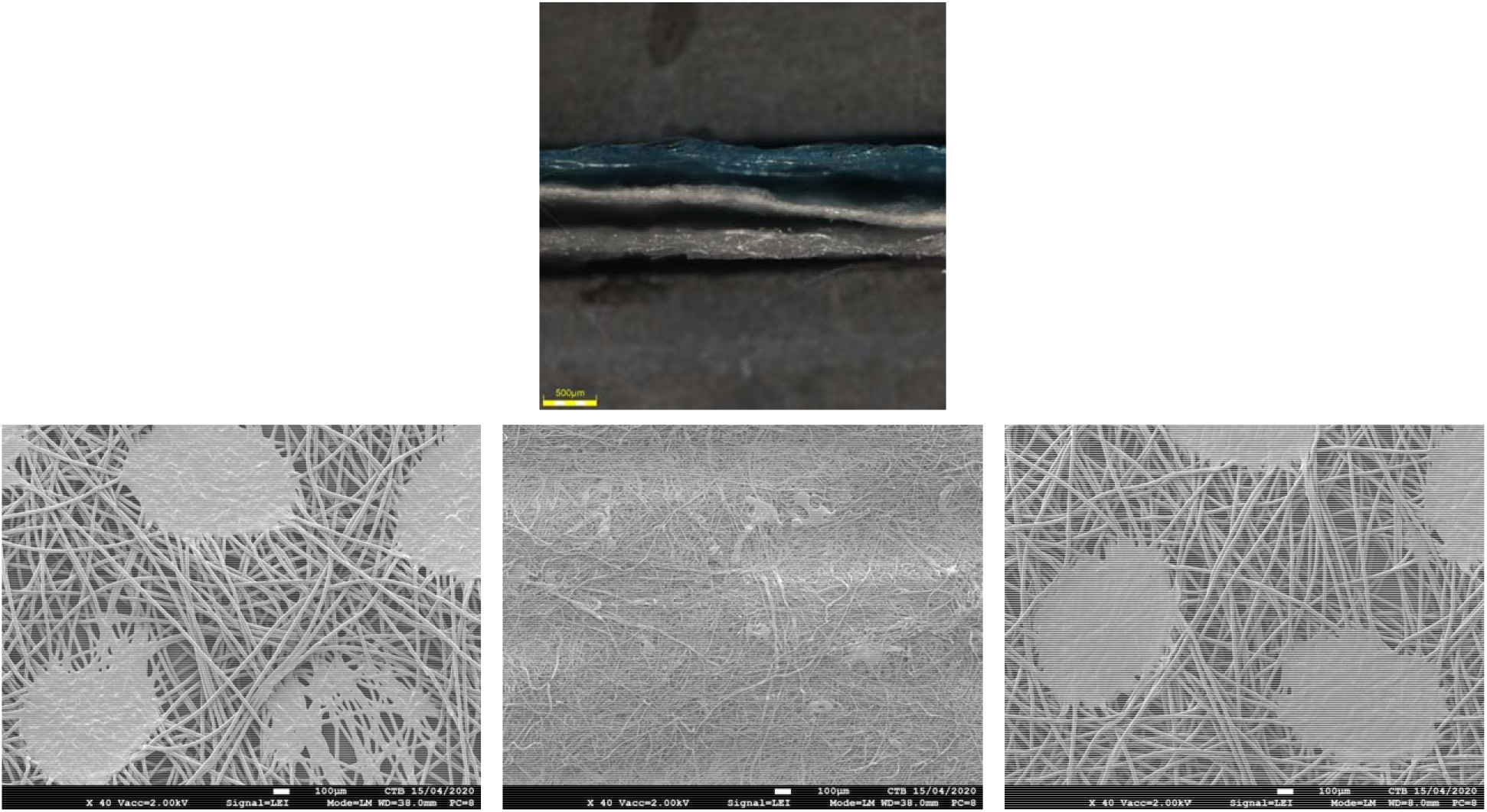
Scanning electron microscopy of a SuninCare™, Protect Plus surgical mask. Top: Cross section view of the surgical mask. The outer layer (blue) is a polypropylene spunbound structure, the middle layer is a polypropylene meltblown structure (26.8 grams/m2) and the inner layer is a polypropylene spunbound structure. Bottom: The outer, intermediate and inner surgical mask layers are depicted from left to right. The dots in the outer and inner spunbound layers show points where the polypropylene fibres are melted together to ensure layer integrity. Pictures were taken on a JEOL JSM-7600F Analytical Ultrahigh resolution thermally assisted field-emission gun source scanning electron microscope at 40 times magnification. Prior to taking the pictures, samples were coated with a platinum/palladium layer (suited to non-conductive samples) using a JEOL JFC-2300 High Resolution sputter coater.

